# Medication use patterns among older patients in temporary stays in Denmark

**DOI:** 10.1101/2024.10.10.24315220

**Authors:** Hanin Harbi, Carina Lundby, Peter Bjødstrup Jensen, Søren Post Larsen, Linda Grouleff Rørbæk, Lene Vestergaard Ravn-Nielsen, Jesper Ryg, Mette Reilev, Kasper Edwards, Anton Pottegård

**Author notes:** Correspondence: Anton Pottegård, Clinical Pharmacology, Pharmacy and Environmental Medicine, Department of Public Health, University of Southern Denmark, Campusvej 55, 5230 Odense M, Denmark.

## Abstract

**Background:** Patients in temporary stays are typically older individuals with frailty and multimorbidity. However, limited knowledge exists about their medication use. This study aimed to describe prescription drug use among patients in temporary stays in Denmark.

**Methods:** We conducted a drug utilisation study on 11,424 patients in public healthcare-operated temporary stay units across 14 Danish municipalities between 2016 to 2023 (median age 81 years; 54% women). Prescription data were sourced from the Danish National Prescription Registry.

**Results:** Patients used a median of 6 drug classes (interquartile range [IQR] 4-10) in the four months before moving into a temporary stay facility; 68% used ≥ 5 drug classes, and 26% used ≥ 10. The most commonly used drug classes were paracetamol (49%), statins (30%), and proton pump inhibitors (29%). The monthly rate of new drug use increased from 23/100 patients six months before move-in to a peak of 262/100 patients in the first month after move-in, driven primarily by laxatives, analgesics, and antibiotics. High-risk drug use increased from 70% to 83% following move-in, with 49% of patients initiating at least one new high-risk drug, most commonly opioids (28%), potassium (17%), and anticoagulants (15%). General practitioners initiated 60%-70% of treatments and maintained 80%-90%. Hospital physician prescriptions increased around move-in, peaking at 55% for initiation and 25% for maintenance in the first month after move-in.

**Conclusion:** Patients in temporary stays in Denmark demonstrate high medication use, including high-risk drugs, with a notable increase in treatment initiations around the time of move-in.

## Introduction

Healthcare systems worldwide are increasingly challenged by ageing populations and the growing burden of chronic diseases. Consequently, patients are being discharged from hospitals earlier, often in less stable condition. This shift has increased the demand for community-based care services to support earlier hospital discharges and prevent hospital (re)admissions [1–5]. One such service is temporary stay facilities, which provide short-term care outside the home, typically for patients who need additional support to recover and regain their strength after a hospital stay. These patients are often older individuals with frailty and multimorbidity. Medication use is substantial among these patients [6–11]. A recent study described the challenges of transitioning patients from hospital to temporary stay facilities, reporting a median use of 8 drugs per patient, with 96% using at least one high-risk drug [12].

Managing medications in temporary stay facilities poses several challenges for care staff. First, care staff do not have access to patients’ medical records. When patients are discharged from a hospital to a temporary stay facility, the care staff does only receive a discharge notice from the hospital nurses, rather than a detailed discharge letter from the hospital physician. Additionally, temporary stay facilities typically do not store medication. Instead, patients must either bring their own medications or have them delivered by a relative or pharmacy. These challenges force care staff to invest considerable time and resources in gathering information about patients’ medication and resolving any discrepancies [8,12].

One study found that only half of the patients discharged from hospitals to temporary stay facilities arrived with all the necessary medication. As a result, nurses had to contact hospital physicians and general practitioners (GPs) repeatedly to clarify medication regimens. It was estimated that one-third of these contacts could have been avoided if the discharge letter had been provided [12].

To improve the treatment and safety of patients in temporary stays, a better understanding is needed not only of medication management challenges but also the quantity and types of medications being used, as well as any changes in medication use around the time of moving into a facility. Therefore, the aim of this study was to describe the use of prescription drugs among patients in temporary stays in Denmark.

## Methods

We established a cohort of 11,424 patients who stayed in temporary care facilities across 14 Danish municipalities during 2016 and 2023. The cohort was supplemented with individual-level data from the Danish National Prescription Registry. The same cohort has been used to describe patient characteristics and care trajectories, which has been published as a preprint [13].

### Data sources

Data on temporary stays were provided by the municipalities for all or part of the study period from January 1, 2016, to December 31, 2023. These data included each patient’s Central Person Register (CPR) number, move-in date, and move-out date. We linked this information to the Danish National Prescription Registry to obtain prescription drug use data. Individual-level linkage was achieved using the CPR number, a unique personal identifier assigned to all Danish residents by the Civil Registration System since 1968 [14].

The Danish National Prescription Registry includes data on all prescription drugs dispensed at Danish community pharmacies since 1995, categorized by the Anatomical Therapeutic Chemical (ATC) classification system. It also contains the dispensing date and prescriber identifier [15]. The prescriber identifier allowed us to categorise prescribers as GPs, private practicing specialists, or hospital physicians. The validity of prescriber data in the Danish National Prescription Registry is high and improving over time [16]. To identify the medical speciality of primary sector prescribers (GP or private practicing specialist), we linked the prescriber identifier to the Registry of Health Care Providers [17]. Data on patient death were obtained from the Civil Registration System [14].

### Study cohort

We included temporary stays if both the move-in and move-out dates occurred within the study period. Temporary stays with missing or invalid CPR numbers, move-in dates, or move-out dates were excluded, as were cases where the move-out date preceded the move-in date. We required patients to have resided in Denmark for at least two years prior to their first temporary stay. For patients with multiple temporary stays, we combined overlapping temporary stays into a single continuous stay. Temporary stays were considered overlapping if there was no gap between the move-out date of one stay and the move-in date of the next. Only the first temporary stay for each patient was included in the analyses.

### Setting

In Denmark, temporary stays are provided by municipalities for individuals requiring short-term care and support that cannot be managed at home. These stays can be used for recovery or rehabilitation following an illness or hospitalisation, as well as for providing respite care for family caregivers. The services and organization of temporary stay facilities vary across municipalities but may involve care from nurses, care assistants, physiotherapists, and occupational therapists. Temporary stay facilities do not typically have physicians on staff, and the primary medical responsibility lies with the patient’s GP or the hospital [8].

### Analyses

First, we described baseline medication use as of the day the patient moved into the temporary stay facility. For each patient, we calculated the median number of drug classes (ATC level 4) dispensed in the four months prior to move-in. We also determined the proportion of patients using at least five drug classes (polypharmacy) and at least ten drug classes (excessive polypharmacy), defined as filling at least one prescription for five or ten different drug classes, respectively, within the four-month period before move-in. Additionally, we identified the most frequently dispensed drug classes and calculated the proportion of patients filling at least one prescription from each main drug group, categorized by the first level of the ATC code (i.e., by target organ or system).

Second, we analysed changes in medication use around the time of moving into the temporary stay facility, focusing on the two years before and after move-in. We calculated the monthly rate of incident drug use per 100 patients. Incident use was defined as the first filled prescription for a drug class that had not been dispensed to the patient within the previous two years. We also identified the drug classes contributing to peaks in incident drug use.

Third, we assessed high-risk drug use at baseline. High-risk drugs were defined according to the Danish Patient Safety Authority and included anticoagulants (ATC code B01A), antidiabetics (A10), digoxin (C01AA05), low-dose methotrexate (L04AX03), opioids (N02A and R05DA04), and potassium (A12B). These drugs are frequently associated with adverse drug events resulting from medication errors and require special attention from healthcare professionals [18]. We excluded gentamicin, which is typically administered intravenously in hospital. For each high-risk drug, we calculated the proportion of patients filling at least one prescription in the following periods: 1) four months before move-in, 2) four months after move-in, 3) before but not after, 4) after but not before, and 5) both periods.

Finally, we described the types of prescribers responsible for initiating treatment (incident prescriptions) and maintaining treatment (nonincident prescriptions). For each month in the two years before and after move-in, we calculated the proportion of incident and nonincident prescriptions issued by GPs, private practicing specialists, and hospital physicians.

All analyses were performed using R version 4.3.3.

### Data and ethics

This study was registered at the University of Southern Denmark’s research inventory (record no. 11.436). Ethical approval is not required for registry-based studies in Denmark.

## Results

We identified 11,424 patients who moved into a temporary stay facility during the study period, analysing only their first temporary stay. The median age of the patients at move-in was 81 years (interquartile range [IQR] 73-87 years), and 54% were women. The median Charlson Comorbidity Index score was 1 (IQR 0-2), and the median number of hospital admissions in the year prior to move-in was 3 (IQR 2-6) (Supplementary Table 1).

The patients used a median of 6 drug classes (IQR 4-10) in the four months prior to moving into the temporary stay facility. Polypharmacy (defined as using at least five drug classes) was observed in 68% of patients, while 26% of patients exhibited excessive polypharmacy (i.e., using at least ten drug classes). The most frequently used drug classes included paracetamol (49%), statins (30%), proton pump inhibitors (29%), platelet inhibitors (26%), and selective beta blockers (23%) (Table 1). The most commonly used drug groups were those related to the nervous system (72%), cardiovascular system (71%), alimentary tract and metabolism (59%), and blood and blood-forming organs (51%) (Supplementary Table 2).

**Table 1.** The 25 most frequently dispensed drug classes at time of moving into a temporary stay facility.

| ATC code | Drug class | Proportion of prevalent users, n (%)<br>(n = 11,424) |
| --- | --- | --- |
| N02BE | Anilides | 5,588 (49) |
| C10AA | HMG CoA reductase inhibitors | 3,463 (30) |
| A02BC | Proton pump inhibitors | 3,274 (29) |
| B01AC | Platelet aggregation inhibitors excl. heparin | 2,995 (26) |
| C07AB | Beta blocking agents, selective | 2,628 (23) |
| C03CA | Sulfonamides, plain | 2,582 (23) |
| A12BA | Potassium | 2,478 (22) |
| C08CA | Dihydropyridine derivatives | 2,173 (19) |
| N02AA | Natural opium alkaloids | 2,001 (18) |
| B01AF | Direct factor Xa inhibitors | 1,752 (15) |
| J01CA | Penicillins with extended spectrum | 1,744 (15) |
| C09AA | ACE inhibitors, plain | 1,652 (14) |
| C09CA | Angiotensin II receptor blockers (ARBs), plain | 1,576 (14) |
| N06AB | Selective serotonin reuptake inhibitors | 1,389 (12) |
| N02AX | Other opioids | 1,292 (11) |
| N06AX | Other antidepressants | 1,177 (10) |
| A06AD | Osmotically acting laxatives | 1,160 (10) |
| A10BA | Biguanides | 1,076 (9.4) |
| R03AC | Selective beta-2 adrenoceptor agonists | 1,056 (9.2) |
| H02AB | Glucocorticoids | 1,054 (9.2) |
| N05CF | Benzodiazepine related drugs | 1,045 (9.1) |
| N02BF | Gabapentinoids | 1,001 (8.8) |
| C03AB | Thiazides and potassium in combination | 856 (7.5) |
| M01AE | Propionic acid derivatives | 843 (7.4) |
| J01CE | Beta-lactamase sensitive penicillins | 843 (7.4) |

<sup>a</sup> Medication use at the day of moving into the temporary stay facility was determined by assessing filled prescriptions within four months prior to move-in.

The monthly rate of incident use increased from 23 per 100 patients six months prior to move-in to 61 per 100 patients in the month before move-in. The rate peaked at 262 per 100 patients per month in the month following move-in, then gradually decreased to a level slightly higher than premove-in around nine months after the move (Figure 1). The primary drug classes responsible for this peak in incidence use were laxatives, analgesics, and antibiotics (Supplementary Table 3, 4, and 5).

**Figure 1.**
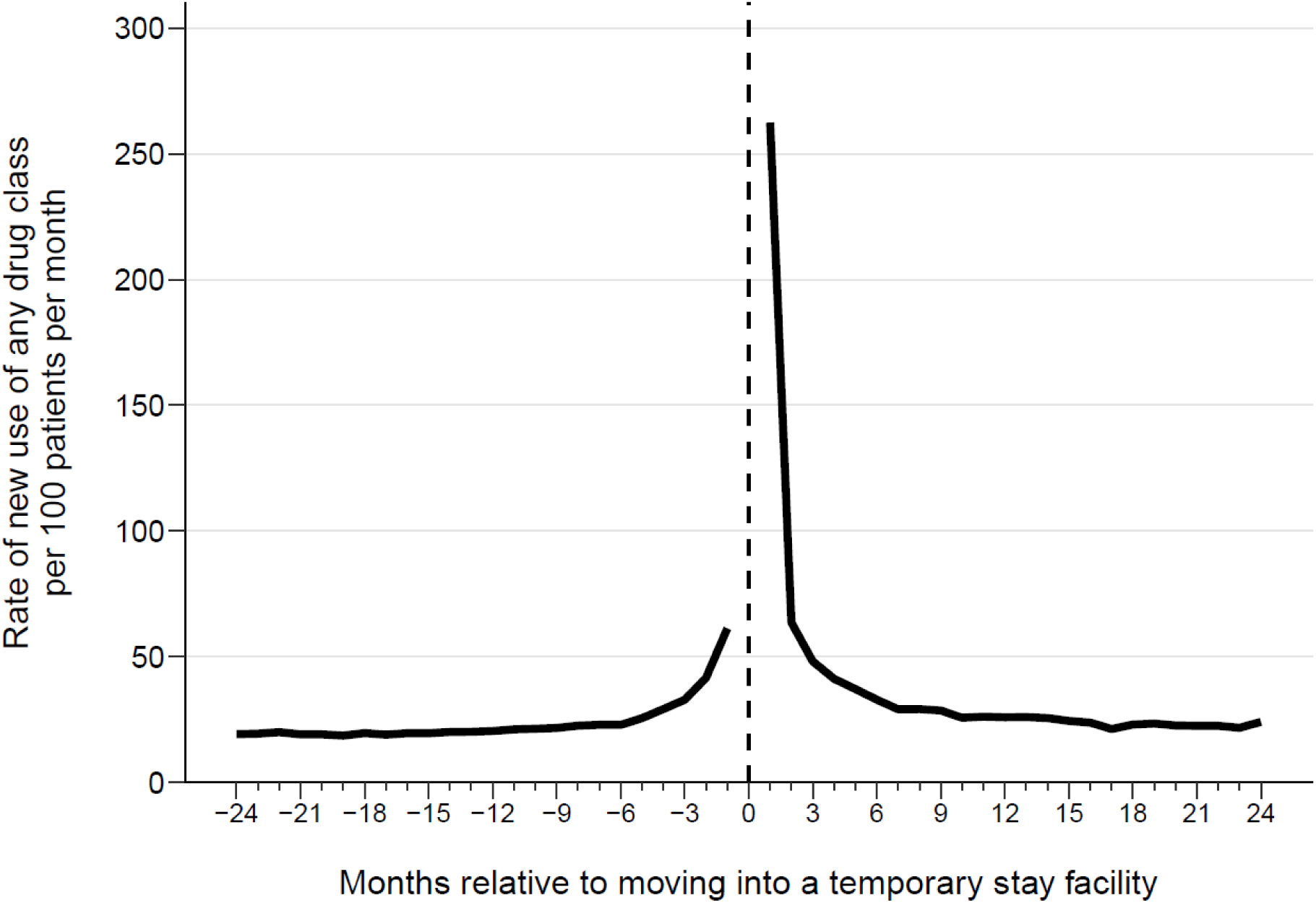
Monthly rate of incident drug use per 100 patients in the two years before and after moving into a temporary stay facility

The proportion of patients using at least one high-risk drug increased from 70% in the four months before move-in to 83% in the four months following move-in. In both periods, the most frequently used high-risk drugs were anticoagulants (46% before, 49% after), opioids (30% before, 51% after), and potassium (22% before, 31% after). Almost half of the patients (49%) used at least one high-risk drug in the four months following move-in that they had not used in the four months prior. The most frequently initiated high-risk drug were opioids (28%), potassium (17%), and anticoagulants (15%). In contrast, 24% of patients used at least one high-risk drug in the four months before move-in that they no longer used in the four months after (Supplementary Table 6). We found similar results when the analysis was restricted to patients who survived the four months following move-in (data not shown).

GPs were responsible for most treatment initiations six months prior to move-in (62%), followed by hospital physicians (24%) and private practicing specialists (6.5%). This pattern remained stable until six months prior to move-in when hospital physicians began issuing a growing share of incident prescriptions, while GPs and private practicing specialists saw a decrease. In the month following move-in, hospital physicians were responsible for the majority of incident prescriptions (55%), with GPs accounting for 39%. By two months after move-in, the distribution of prescriber types had returned to levels similar to those before move-in, although with a slight increase in the proportion of incident prescriptions issued by GPs and a decrease in those issued by hospital physicians and private practicing specialists (Figure 2).

**Figure 2.**
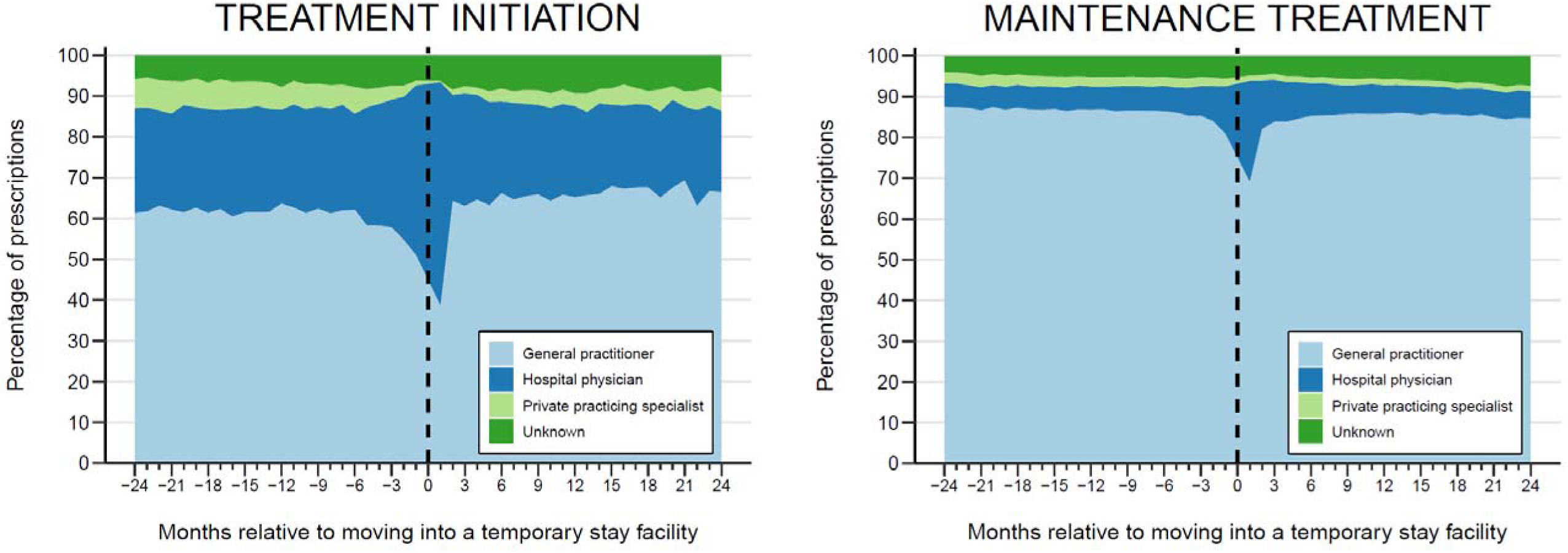
Monthly distribution of prescriber types responsible for initiating treatments (incident prescriptions, left panel) and maintaining treatments (nonincident prescriptions, right panel) in the two years before and after moving into a temporary stay facility.

For maintenance treatment, GPs issued 80-90% of nonincident prescriptions, though this proportion dropped to 69% in the month after move-in. During this period, hospital physicians’ share of maintenance prescriptions rose to 25%, while private practicing specialists’ share decreased to 1.2%.

## Discussion

In this drug utilisation study, we describe medication use among patients in temporary stays in Denmark. We found a high level of polypharmacy, with half of the patients using at least six drug classes at the time of move-in. Treatment initiations increased sharply around move-in and remained slightly elevated thereafter. Patients had a substantial use of high-risk drugs, which increased after move-in. GPs were the primary prescribers for both initiating and maintaining treatments, although hospital physicians played a crucial role in initiating new treatments around move-in.

The primary strength of this study is the use of a nationwide registry with high-quality prescription fill data [15,19], ensuring comprehensive patient coverage and reducing the potential for selection bias. An additional advantage is that the registry captures filled rather than issued prescriptions, eliminating the impact of primary nonadherence, when patients fail to fill an initial prescription for a new drug [20].

A limitation of the study is that it relies on prescription fill data, so we cannot be certain that patients took the drugs as prescribed. However, a recent Danish study found that most patients (68%) who moved into temporary stays after hospital discharge received help with medication management before hospitalization [12]. Additionally, healthcare staff at Danish temporary stay facilities provide support with medication management [8], minimizing the risk of nonadherence impacting on our results. Another limitation is that some drugs can be purchased over the counter, which are not recorded in the Danish National Prescription Registry [15]. However, as many patients in temporary stay receive help with medication management, which typically involves prescribed medications, this limitation may be mitigated. Finally, because the Danish National Prescription Registry does not capture drugs used during hospital stays [15], medications initiated during hospital admissions may appear as new prescriptions after move-in, potentially inflating the initiation rate following move-in.

This study is the first to systematically describe medication use among patients in temporary stays across Denmark. A recent study on a smaller cohort of patients moving into a temporary stay facility after hospital discharge reported a median of 8 drugs at move-in [12]. This is higher than the median in our study, possibly due to differences in the populations studied (a single temporary stay facility versus nationwide data) or because their results were based on issued rather than filled prescriptions. This may also explain their higher reported use of opioids (47% versus 30%) and digoxin (14% versus 5.6%). The substantially increased use of high-risk drugs reported in their study (96% versus 70%) is likely explained by their broader definition of risk drugs, which included frequently used medications such as beta blockers and angiotensin-converting enzyme (ACE) inhibitors or angiotensin II receptor blockers. To our knowledge, only few international studies have described patients in temporary stays, none of which provide detailed information about their medication use [21–24].

We observed a marked increase in treatment initiations around move-in, with laxatives, analgesics, and antibiotics being the most frequently initiated drug classes. This finding aligns with previous research on Danish care home residents [25], suggesting that the health decline leading to a temporary stay often results in increased medical interventions, including hospitalisation and GP visits. Increased medical attention likely contributes to medication changes, including the initiation of drugs previously purchased over the counter, such as laxatives. Notably, 86% of patients moving into a temporary stay facility after hospital discharge have at least one drug initiated during their hospital stay [12]. Although we observed a substantial increase in prescriptions initiated by hospital physicians around move-in, GPs continued to play a considerable role in initiating new treatments [26].

Medication use among patients in temporary stays closely mirrors that of Danish care home residents [25]. Both populations tend to be older, with multimorbidity and polypharmacy, which increases their risk of adverse drug effects and interactions. While certain drugs, such as opioids (frequently initiated around move-in), are necessary for symptom management, others, such as cholesterol-lowering drugs, may offer limited benefit. For older patients with frailty and limited life expectancy, the time to benefit from some medications may exceed their remaining lifespan [27,28]. This underscores the importance of regular medication reviews to align treatment plans with the patient’s current health status and care goals.

Patients in temporary stays are particularly vulnerable to adverse drug events due to medication errors, which can lead to serious consequences. Ensuring smooth transitions and clarity around medical management is critical for these patients. An example of an effort to improve medication safety is Denmark’s recently implemented “72-hour extended treatment responsibility”, where discharging hospital departments remain responsible for a patient’s treatment for 72 hours after discharge to municipal care. During this period, municipal healthcare professionals can consult the discharging hospital for medical advice or guidance. Evaluations of this agreement highlight its success in strengthening communication and improving medication safety, with most calls coming from temporary stay facilities and concerning medication-related issues [29,30].

In conclusion, we found a high level of medication use among patients in temporary stays, with a marked increase in new treatment initiations around move-in. The use of high-risk drugs also increased after move-in, underscoring the need for careful medication management in these settings. Our findings provide valuable insights that can help guide efforts to optimize medication use and management in temporary stay facilities, ultimately improving patient safety and quality of life.

## Data Availability

The data was used under license from the Danish Health Data Authority.

---

**Supplementary Table 1.** Baseline characteristics of patients moving into temporary stay facilities in 14 Danish municipalities from 2016 to 2023.

|  | Total<br>(n = 11,424) |
| --- | --- |
| Sex |  |
| Female | 6,141 (54%) |
| Male | 5,283 (46%) |
| Age |  |
| Median (IQR) | 81 (73-87) |
| < 75 years | 3,394 (30%) |
| 75-84 years | 4,017 (35%) |
| ≥ 85 years | 4,013 (35%) |
| Charlson Comorbidity Index (CCI) |  |
| Median (IQR) | 1 (0-2) |
| 0-1 | 5,752 (50%) |
| 2-3 | 3,921 (34%) |
| ≥ 4 | 1,751 (15%) |
| Hospitalizations in the year before move-in |  |
| Median (IQR) | 3 (2-6) |
| 0-2 | 4,209 (37%) |
| 3-5 | 3,939 (34%) |
| ≥ 6 | 3,276 (29%) |

**Supplementary Table 2.** Use of main drug groups, defined by the first level of the Anatomical Therapeutic Chemical (ATC) code, at time of moving into a temporary stay facility.

| ATC code | Main drug group | Proportion of prevalent users, n (%)<br>(n = 11,424) |
| --- | --- | --- |
| N | Nervous system | 8,240 (72) |
| C | Cardiovascular system | 8,077 (71) |
| A | Alimentary tract and metabolism | 6,702 (59) |
| B | Blood and blood-forming organs | 5,779 (51) |
| J | Antiinfectives for systemic use | 3,848 (34) |
| R | Respiratory system | 2,797 (24) |
| M | Musculoskeletal system | 2,477 (22) |
| H | Systemic hormonal preparations, excl. sex hormones and insulins | 1,881 (16) |
| G | Genito urinary system and sex hormones | 1,823 (16) |
| D | Dermatologicals | 1,498 (13) |
| S | Sensory organs | 1,479 (13) |
| P | Antiparasitic products, insecticides and repellents | 333 (2.9) |
| L | Antineoplastic and immunomodulating agents | 152 (1.3) |
| V | Various | 24 (0.21) |

<sup>a</sup> Medication use at the day of moving into the temporary stay facility was determined by assessing filled prescriptions within four months prior to move-in.

**Supplementary Table 3.**
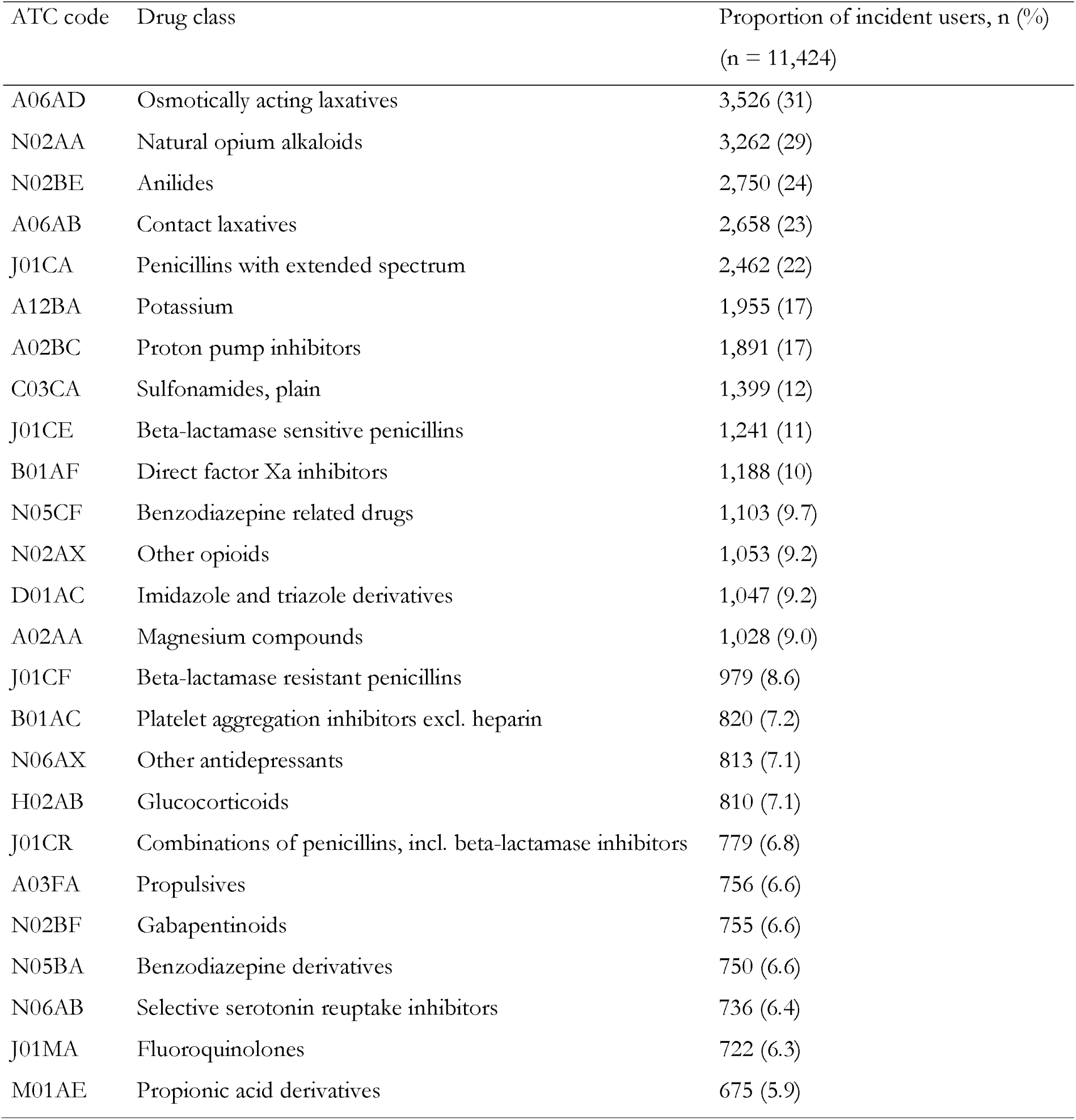
Top 25 most frequently initiated drug classes during the peak incidence rate observed around move-in (i.e., from five months before to four months after move-in)

**Supplementary Table 4.** Top 25 most frequently initiated drug classes during the peak incidence rate in the premove-in phase (i.e., the five months before move-in)

| ATC code | Drug class | Proportion of incident users, n (%)<br>(n = 11,424) |
| --- | --- | --- |
| N02AA | Natural opium alkaloids | 1,090 (9.5) |
| J01CA | Penicillins with extended spectrum | 909 (8.0) |
| N02BE | Anilides | 899 (7.9) |
| A06AD | Osmotically acting laxatives | 775 (6.8) |
| A12BA | Potassium | 633 (5.5) |
| A02BC | Proton pump inhibitors | 620 (5.4) |
| J01CE | Beta-lactamase sensitive penicillins | 614 (5.4) |
| A06AB | Contact laxatives | 548 (4.8) |
| N02AX | Other opioids | 537 (4.7) |
| C03CA | Sulfonamides, plain | 488 (4.3) |
| H02AB | Glucocorticoids | 425 (3.7) |
| J01CF | Beta-lactamase resistant penicillins | 412 (3.6) |
| M01AE | Propionic acid derivatives | 404 (3.5) |
| B01AF | Direct factor Xa inhibitors | 367 (3.2) |
| N02BF | Gabapentinoids | 338 (3.0) |
| J01CR | Combinations of penicillins, incl. beta-lactamase inhibitors | 336 (2.9) |
| D01AC | Imidazole and triazole derivatives | 333 (2.9) |
| A02AA | Magnesium compounds | 325 (2.8) |
| N05CF | Benzodiazepine related drugs | 308 (2.7) |
| A03FA | Propulsives | 304 (2.7) |
| N06AX | Other antidepressants | 290 (2.5) |
| J01MA | Fluoroquinolones | 286 (2.5) |
| N06AB | Selective serotonin reuptake inhibitors | 283 (2.5) |
| B01AC | Platelet aggregation inhibitors excl. heparin | 251 (2.2) |
| N05BA | Benzodiazepine derivatives | 232 (2.0) |

**Supplementary Table 5.**
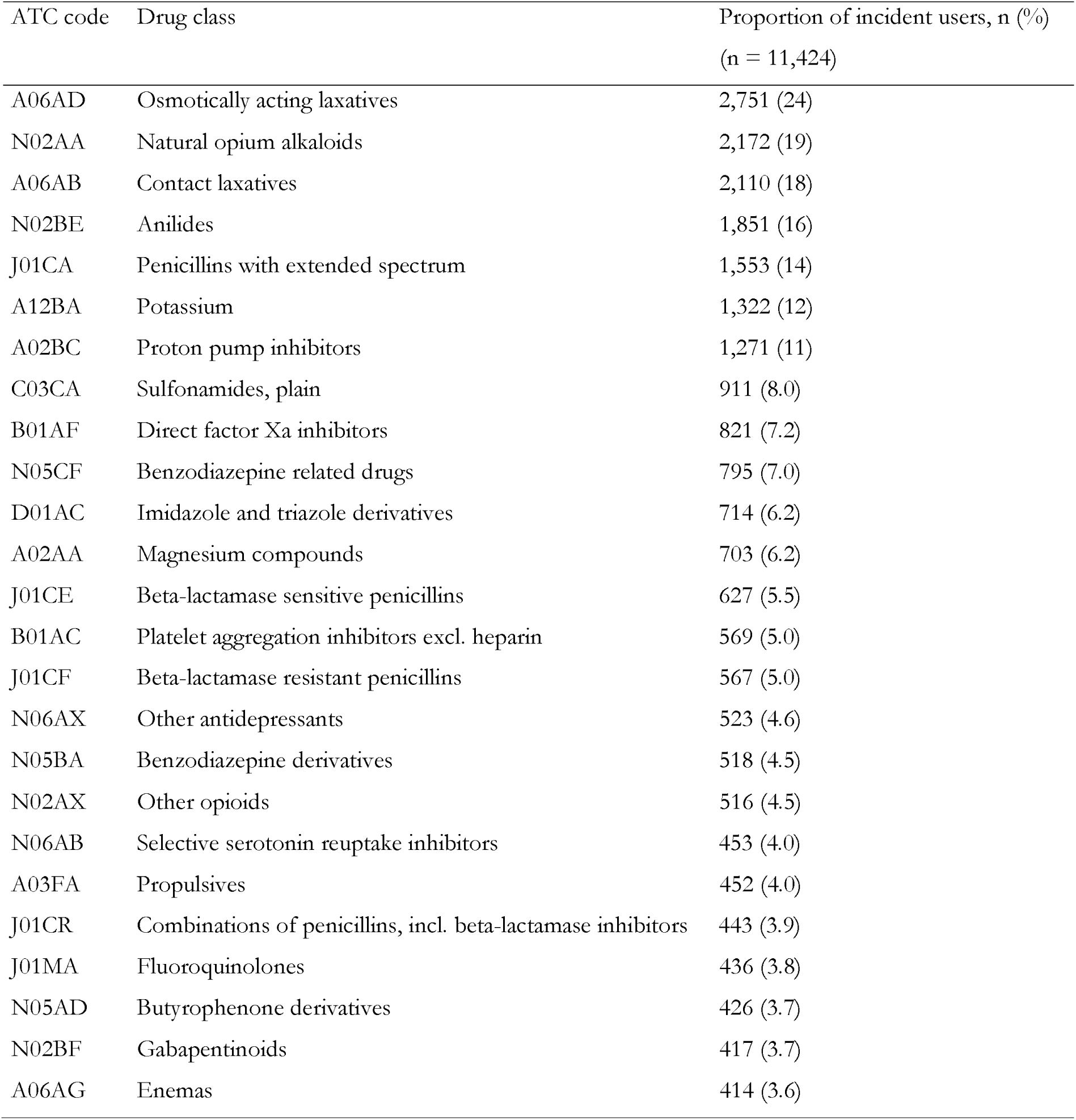
Top 25 most frequently initiated drug classes during the peak incidence rate in the postmove-in phase (i.e., the four months after move-in)

**Supplementary Table 6.** Proportion of patients (n = 11,424) filling at least one prescription for a high-risk drug in the four months before and after moving into a temporary stay facility.

|  | 4 months before move-in,<br>n (%) | 4 months after move-in,<br>n (%) | Only during the 4<br>months before move-in,<br>n (%) | Only during the 4<br>months after move-in,<br>n (%) | During both the 4 months<br>before and after move-in,<br>n (%) |
| --- | --- | --- | --- | --- | --- |
| Anticoagulants | 5,245 (46) | 5,627 (49) | 1,333 (12) | 1,715 (15) | 3,912 (34) |
| Antidiabetics | 1,831 (16) | 1,760 (15) | 374 (3.3) | 303 (2.7) | 1,457 (13) |
| Digoxin | 641 (5.6) | 750 (6.6) | 210 (1.8) | 319 (2.8) | 431 (3.8) |
| Low-dose methotrexate | 80 (0.70) | 68 (0.60) | 37 (0.32) | 25 (0.22) | 43 (0.38) |
| Opioids | 3,425 (30) | 5,794 (51) | 804 (7.0) | 3,173 (28) | 2,621 (23) |
| Potassium | 2,466 (22) | 3,523 (31) | 851 (7.4) | 1,908 (17) | 1,615 (14) |
| Any | 7,949 (70) | 9,434 (83) | 2,729 (24) | 5,612 (49) | 6,600 (58) |

## References

1. WHO (World Health Organization). 2015. World report on aging and health. Accessed October 3, 2024. https://iris.who.int/bitstream/handle/10665/186463/9789240694811_eng.pdf?sequence=1.

2. Mac Arthur I, Hendry A. Inter-American Development Bank. 2017. The “Intermediate Care Hospital”: Facility Bed-Based Rehabilitation for Elderly Patients. Accessed October 4, 2024. 10.18235/0009360.

3. Sezgin D, O’Caoimh R, O’Donovan MR et al. Defining the characteristics of intermediate care models including transitional care: an international Delphi study. Aging Clin Exp Res 2020;32:2399–410.

4. Sezgin D, O’Caoimh R, Liew A et al. The effectiveness of intermediate care including transitional care interventions on function, healthcare utilisation and costs: a scoping review. Eur Geriatr Med 2020;11:961–74.

5. Griffiths PD, Edwards ME, Forbes A et al. Effectiveness of intermediate care in nursingl_Jled inl_Jpatient units. Cochrane Database Syst Rev 2007;2007:CD002214.

6. Martinsen B, Norlyk A, Lomborg K. Experiences of intermediate care among older people: a phenomenological study. Br J Community Nurs 2015;20:74–9.

7. The Danish Organization of General Practitioners and Local Government Denmark. [Definition of temporary 24-hour placements] (in Danish). Accessed August 22, 2024. https://stps.dk/Media/638257904106148719/plo_og_kl_-_definition_af_midlertidige_d-gnd-kkede.pdf.

8. Vinge S, Buch MS, Kjellberg PK. The Danish Center for Social Science Research. 2021. [The municipal emergency area - Experiences and perspectives on the development from 15 municipalities] (in Danish). Accessed August 22, 2024. https://www.vive.dk/media/pure/16209/5709970.

9. Danish Ministry of Health and Senior Citizens. 2016. [Enhanced effort for the older medical patient - National action plan 2016] (in Danish). Accessed August 22, 2024. https://sm.dk/Media/637666125515729596/AE_Styrket_indsats_for_den_aeldre_medicinske_patient_national_handlingsplan_2016.pdf.

10. Kayser K, Christensen MR. Municipality of Copenhagen. 2018. [2. discussion of strategy for temporary stays] (in Danish). Accessed August 22, 2024. https://www.kk.dk/dagsordener-og-referater/Sundheds-%20og%20Omsorgsudvalget/m%C3%B8de-24052018/referat/punkt-7.

11. Kayser K, Christensen MR. Municipality of Copenhagen. 2021. [Status regarding temporary stays] (in Danish). Accessed August 22, 2024. https://www.kk.dk/dagsordener-og-referater/Sundheds-%20og%20Omsorgsudvalget/m%C3%B8de-22062021/referat/punkt-3.

12. Ravn-Nielsen LV, Bjørk E, Nielsen M et al. Challenges related to transitioning from hospital to temporary care at a skilled nursing facility: a descriptive study. Eur Geriatr Med 2024, DOI: 10.1007/s41999-024-01003-z.

13. Harbi H, Lundby C, Jensen PB et al. Characteristics and care trajectories of older patients in temporary stays in Denmark. Preprint made available at medrxiv.com.

14. Schmidt M, Pedersen L, Sørensen HT. The Danish Civil Registration System as a tool in epidemiology. Eur J Epidemiol 2014;29:541–9.

15. Pottegård A, Schmidt SAJ, Wallach-Kildemoes H et al. Data Resource Profile: The Danish National Prescription Registry. Int J Epidemiol 2017;46:798–798f.

16. Rasmussen L, Valentin J, Gesser KM et al. Validity of the Prescriber Information in the Danish National Prescription Registry. Basic Clin Pharmacol Toxicol 2016;119:376–80.

17. The Danish Health Data Authority. [The Registry of Health Care Providers] (in Danish). Accessed August 22, 2024. https://sundhedsdatastyrelsen.dk/da/registre-og-services/om-de-nationale-sundhedsregistre/personoplysninger-og-sundhedsfaglig-beskaeftigelse/yderregisteret.

18. The Danish Patient Safety Authority. [High-risk drugs] (in Danish). Accessed August 22, 2024. https://stps.dk/sundhedsfaglig/viola-viden-og-laering/risikoomraader/risikosituationslaegemidler.

19. Harbi H, Pottegård A. Validation of the “Indication for Use” (INDO) Variable in the Danish National Prescription Registry. Epidemiology 2024;35:1–6.

20. Pottegård A, Christensen R dePont, Houji A et al. Primary non-adherence in general practice: a Danish register study. Eur J Clin Pharmacol 2014;70:757–63.

21. Evans CJ, Potts L, Dalrymple U et al. Characteristics and mortality rates among patients requiring intermediate care: a national cohort study using linked databases. BMC Med 2021;19:48.

22. Nystrøm V, Lurås H, Moger T et al. Finding good alternatives to hospitalisation: a data register study in five municipal acute wards in Norway. BMC Health Serv Res 2022;22:715.

23. Nilsen H, Hunskaar S, Ruths S. Patient trajectories in a Norwegian unit of municipal emergency beds. Scand J Prim Health Care 2017;35:137–42.

24. Schmidt AK, Lilleeng B, Baste V et al. First four years of operation of a municipal acute bed unit in rural Norway. Scand J Prim Health Care 2018;36:390–6.

25. Lundby C, Jensen J, Larsen SP et al. Use of medication among nursing home residents: a Danish drug utilisation study. Age Ageing 2020;49:814–20.

26. Pottegård A, Olesen M, Christensen B et al. Who prescribes drugs to patients: A Danish register-based study. Br J Clin Pharmacol 2021;87:2982–7.

27. Holmes HM, Hayley DC, Alexander GC et al. Reconsidering medication appropriateness for patients late in life. Arch Intern Med 2006;166:605–9.

28. Poudel A, Yates P, Rowett D et al. Use of Preventive Medication in Patients With Limited Life Expectancy: A Systematic Review. J Pain Symptom Manage 2017;53:1097–1110.e1.

29. Center for Clinical Research and Prevention, Frederiksberg, The Capital Region of Denmark. 2023. [Evaluation of 72-hour extended treatment responsibility - Interim report 1] (in Danish). Accessed September 26, 2024. https://www.regionh.dk/til-fagfolk/Sundhed/Tvaersektorielt-samarbejde/Documents/Evaluering%20af%20aftalen%20om%2072%20timers%20behandlingsansvar%20-%20delrapport%201.pdf.

30. Center for Clinical Research and Prevention, Frederiksberg, The Capital Region of Denmark. 2024. [Evaluation of 72-hour extended treatment responsibility - Interim report 2] (in Danish). Accessed September 26, 2024. https://www.regionh.dk/til-fagfolk/Sundhed/Tvaersektorielt-samarbejde/Documents/Evaluering%20af%20aftalen%20om%2072%20timers%20behandlingsansvar%20-%20delrapport%202.pdf.

